# The N501Y mutation in SARS-CoV-2 spike leads to morbidity in obese and aged mice and is neutralized by convalescent and post-vaccination human sera

**DOI:** 10.1101/2021.01.19.21249592

**Authors:** Raveen Rathnasinghe, Sonia Jangra, Anastasija Cupic, Carles Martínez-Romero, Lubbertus C.F. Mulder, Thomas Kehrer, Soner Yildiz, Angela Choi, Ignacio Mena, Jana De Vrieze, Sadaf Aslam, Daniel Stadlbauer, David A. Meekins, Chester D. McDowell, Velmurugan Balaraman, Juergen A. Richt, Bruno G. De Geest, Lisa Miorin, PVI study group, Florian Krammer, Viviana Simon, Adolfo García-Sastre, Michael Schotsaert

## Abstract

The current COVID-19 (coronavirus disease 19) pandemic, caused by SARS-CoV-2, disproportionally affects the elderly and people with comorbidities like obesity and associated type 2 diabetes mellitus. Small animal models are crucial for the successful development and validation of antiviral vaccines, therapies and to study the role that comorbidities have on the outcome of viral infections. The initially available SARS-CoV-2 isolates require adaptation in order to use the mouse angiotensin converting enzyme 2 (mACE-2) entry receptor and to productively infect the cells of the murine respiratory tract. We have “mouse-adapted” SARS-CoV-2 by serial passaging a clinical virus isolate in the lungs of mice. We then used low doses of this virus in mouse models for advanced age, diabetes and obesity. Similar to SARS-CoV-2 infection in humans, the outcome of infection with mouse-adapted SARS-CoV-2 resulted in enhanced morbidity in aged and diabetic obese mice. Mutations associated with mouse adaptation occurred in the S, M, N and ORF8 genes. Interestingly, one mutation in the receptor binding domain of the S protein results in the change of an asparagine to tyrosine residue at position 501 (N501Y). This mutation is also present in the newly emerging SARS-CoV-2 variant viruses reported in the U.K. (20B/501Y.V1, B1.1.7 lineage) that is epidemiologically associated with high human to human transmission. We show that human convalescent and post vaccination sera can neutralize the newly emerging N501Y virus variant with similar efficiency as that of the reference USA-WA1/2020 virus, suggesting that current SARS-CoV-2 vaccines will protect against the 20B/501Y.V1 strain.

## Introduction

Severe acute respiratory syndrome coronavirus-2 (SARS-CoV-2) is the cause of the present coronavirus disease 2019 (COVID-19) pandemic which has claimed hundreds of thousands of lives with the death toll still rising. The virus belongs to the family *Coronaviridae* and genera Betacoronaviruses which consists of a single stranded, positive sense ∼30kB RNA as genome. The genome encodes for four structural proteins – nucleoprotein (NP), spike (S), envelope (E) and membrane (M) proteins. The S protein of the virus plays an integral part in viral fusion and entry into host cells. The homotrimeric S protein consists of S1 and S2 subunits. The receptor binding domain (RBD) of the S1 subunit binds to angiotensin-converting enzyme 2 (ACE-2) present on the host cellular surfaces. The interaction is followed by S2-driven-transcleavage of the S protein by cellular metalloproteases such as TMPRSS2, thereby facilitating an efficient fusion and release of viral contents into the host cell (1). Since the RBD domain is necessary for direct interaction with the host receptor, mutations in RBD can affect SARS-CoV-2 infection efficiency depending on the host. As the RBD is also the target of virus-neutralizing antibodies, mutations in RBD can impact the neutralizing titers of polyclonal and monoclonal antibodies.

The virus primarily spreads through respiratory droplets and causes a diverse array of symptoms from completely asymptomatic infections to fever, cough, anosmia, pneumonia, acute respiratory distress syndrome, microvascular coagulation, multi-organ dysfunction and other severe manifestations -including death-in humans. Several predisposition factors have been found to be associated with increased susceptibility to severe COVID-19. Hypertension, cardiovascular diseases, gender, advanced age and obesity have already been defined as major risk factors among humans attributing to increased mortality by enhancing secondary conditions such as hypoxemia and pneumonia (2–7). In order to study SARS-CoV-2-associated comorbidities, there is a need for small animal models in which pulmonary inflammation, lung lesions and histopathology after SARS-CoV-2 infection can be investigated. The RBD of the S protein from the SARS-CoV-2 strain that started the pandemic does not efficiently bind mouse ACE-2 (mACE-2, a murine ortholog of human-ACE-2) (8) and as a consequence this SARS-CoV-2 strain does not efficiently infect laboratory mouse strains efficiently. Several approaches have been developed to allow the use of mouse models for SARS-CoV-2 research (9–11), often guided by the experience obtained previously for mouse models to study SARS-CoV. Many of these models rely on transgenic mice that express hACE-2 in epithelial cells or sensitize mice to SARS-CoV-2 infection by adenovirus-mediated transduction of the hACE-2 gene (Ad-hACE-2) in the respiratory tract (9,12). These models have been crucial for studying host-pathogen interactions, prophylactic and therapeutic interventions in the context of SARS-CoV and SARS-CoV-2 infection. However, a major drawback of these models is that expression of hACE-2 is often driven by a promoter that is not the original ACE-2 promoter, and therefore promoter control and expression patterns can differ and, in the case of Ad-hACE-2, depend on transduction efficiency. These problems would be circumvented by a mouse-adapted SARS-CoV-2 (MA-SARS-CoV-2) that uses the endogenously expressed mACE-2. Moreover, a MA-SARS-CoV-2 can be used with already established mouse models of the comorbidities associated with more severe COVID-19 and allows the efficient exploiting of the genetic toolboxes available for mice.

In this study, we have developed and characterized a MA-SARS-CoV-2 strain after serially passaging a clinical virus isolate (USA-WA1/2020) first in immune-compromised followed by immune-competent mice. We mapped mutations associated with mouse adaptation in the SARS-CoV-2 genome and observed that one of them is the N501Y mutation in the RBD from the spike protein that was also reported for the newly emerging SARS-CoV-2 variant (20B/501Y.V1 strain) with potentially enhanced human transmission potential. This mouse-adapted SARS-CoV-2 strain with N501Y mutation causes enhanced morbidity in mouse models for advanced age, obesity and obesity-associated type 2 diabetes mellitus. Finally, we show that human sera from convalescent and vaccinated individuals can neutralize both the reference USA-WA1/2020 strain and the mouse adapted strain that contains the N501Y spike mutation with similar efficiency.

## Materials and Methods

### Reagents

All chemicals for synthesis were purchased from Thermofisher, unless noted otherwise. Horse radish peroxidase (HRP)-conjugated anti-mouse IgG antibody was purchased from Abcam (ab6823). Anti-mouse SARS-CoV-2 NP (NP1C7C7) and anti-mouse SARS-CoV-2 spike (2BCE5) antibodies were obtained from Center for Therapeutic Antibody Development at Icahn School of Medicine at Mount Sinai, New York. Antibodies used in Western blot were anti-mouse-ACE-2 (R&D, MAB3437), anti-beta-tubulin (Sigma Aldrich, T8328) and anti-mouse (HRP-conjugated, KwikQuant).

### Cell lines

Vero-E6 cells (ATTC-CRL 1586, clone E6) are routinely cultured in the laboratory and were maintained in Dulbecco’s Modified Eagle’s Medium supplemented with 10% fetal bovine serum (FBS, Hyclone), penicillin/streptomycin and 1% non-essential amino acids. Mouse ACE-2 (mACE-2) expressing Vero-E6 cells were established by transducing Vero-E6 cells with a lentiviral vector expressing mACE-2 and a puromycin resistance gene (supplementary figure S1). Cells were selected as a polyclonal population by puromycin selection and mACE-2 expression was confirmed by Western blot.

### Mouse models for comorbidities

All mice strains were obtained from Charles River Laboratories, MA and were housed in a pathogen-free facility at Icahn School of Medicine at Mount Sinai, with food and water ad libitum, adhering to the guidelines from Institutional Animal Care and Use Committee (IACUC). To establish obese mice models, C57Bl6 mice were fed with control or high fat diets (Research Diets). Mice body weights were recorded over 14 weeks followed by diabetic profiling by intraperitoneal glucose tolerance test. Briefly, mice were moved to fresh cages and fasted for 6 hours with access only to water. Mice were then injected intraperitoneally with dextrose solution at 2g/kg body weight. Blood from mice was drawn by sub-mandibular bleed at 0 min, 30 min and 60 min of injection. Blood glucose levels were estimated by Glucose Assay Kit (abcam) by extrapolation from a standard curve. These obese mice were challenged with MA-SARS-CoV-2 to study the severity of disease progression. To study age as a risk factor for SARS-CoV-2-linked disease severity, 6-8-weeks or 52-weeks-old C57Bl6 mice were used for mouse-adapted-SARS-CoV-2 challenge.

### SARS-CoV-2 isolates and mouse adaptation

SARS-CoV-2 isolate USA-WA1/2020 (BEI resources; NR-52281), referred in this manuscript as WT-SARS-CoV-2, was used to challenge mice intranasally. A variant of virus (termed MA-SARS-CoV-2) was obtained after series of passaging in different backgrounds of laboratory mice as well as mACE-2 expressing VeroE6 cells. Briefly, the virus was serially passaged every 2 days via intranasal inoculation of the virus in 50 ul volume derived from the spun-down supernatants of lung homogenates. The mouse adaptation of the SARS-CoV-2 variant was studied in C57Bl6, BALB/c and 129S1/SVMJ (termed 129 for simplicity in the text and figures) mice models. Viral stocks were sequenced after propagation to verify the integrity of the original viral genome.

### Deep sequencing of the viral stocks

To sequence the viral stocks we followed the protocol developed by ARTIC (https://artic.network/ncov-2019) with the primer set version 3. Viral RNA was purified using Viral-RNA kit (mega-Bio-Tek) following the manufacturer instructions and used as template to prepare a cDNA. Overlapping amplicons of ∼400 bp covering the whole genome where barcoded using the Oxford Nanopore Technologies (ONT) Native Barcoding Expansion kit (EXP-NBD104). Libraries where prepared according to the manufacturer instructions, loaded on a minION sequencer equipped with a FLO-MIN106D flow cell. The consensus sequence was obtained using Lasergene software (DNAstar).

### Multi-cycle growth curve for WT and MA-SARS-CoV-2

Confluent Vero-E6 cells in 24 well format were infected with a multiplicity of infection (MOI) of 0.001 of either WT or MA-SARS-CoV-2 virus for 45 minutes, the inoculum was then removed before supplementing with viral growth media (1x Minimal Essential Medium + 2% FBS + 1% penicillin/streptomycin). Each well was considered as one replicate per timepoint and supernatants were stored at –80°C. Viral titers were determined by plaque assay for each sample.

### Virus challenge

2.5⨯10^4^ plaque forming units (PFU) per mice of WT- or MA-SARS-CoV-2 were used for intranasal infection, unless specified otherwise under mild ketamin/xylazine sedation. Body weights were recorded every day to assess the morbidity post-infection until organ harvest. The organs were homogenized in 1x phosphate buffered saline (PBS) and virus titers were determined by plaque assay. Blood for serology or micro-neutralization assays was collected either by submandibular bleeding technique or terminally by cardiac puncture.

### Plaque assay

Plaque assays were performed to determine viral titers in samples or organs harvested from mice challenged with WT or MA-SARS-CoV-2. Briefly, lungs or other organs were harvested from the mice and homogenized in sterile 1X PBS. After brief centrifugation (10,000 g x 5 minutes), the tissue debris was discarded, and the supernatant was 10-fold serially diluted starting from 1:10 dilution. Pre-seeded Vero-E6 or mACE-2-Vero-E6 cells (for WT and MA-SARS-CoV-2 respectively) were infected with tissue homogenate for 1 hour at room temperature (RT) followed by an overlay of 2% Oxoid agar mixed with 2X MEM supplemented with 0.3% FBS. The cells were incubated for 72 hours at 37°C and 5% CO_2_ followed by fixation in 1ml of 4% methanol-free formaldehyde. The plaques were immune-stained with anti-mouse SARS-CoV-2-NP and anti-mouse SARS-CoV-2-spike antibodies for 1 hour at RT and consequently with HRP-conjugated anti-mouse secondary IgG antibody for 1 hour at room temperature (RT). Finally, the plaques were developed with TrueBlue substrate (KPL-Seracare) and viral titers were calculated and expressed as plaque forming units (PFU)/ml.

### Western Blot

Cells were lysed in RIPA buffer (Sigma Aldrich, USA) supplemented with protease inhibitor cocktail (Roche, Switzerland). Total protein concentration was determined in each sample by BCA assay and normalized. The lysates were run on a 4-20 % gradient polyacrylamide gel at 60V and transferred onto polyvinylidene fluoride (PVDF) membranes (BioRad Laboratories) using BIO-RAD semi-dry transfer system. PVDF membranes were blocked in 5 % non-fat dry milk-containing Tris-buffered saline with Tween-20 (TBST) containing 0.1% Tween-20. Anti-Tubulin and anti-mACE-2 (R&D Systems, Cat# MAB3437) primary antibodies were used at dilution of 1:1000 while secondary HRP-conjugated antibodies were used at dilutions of 1:10000 in 3% BSA-containing TBST.

### 50% tissue culture infective dose (TCID_50_) calculation and *in vitro* micro-neutralization assay

To estimate the neutralizing efficiency of sera from vaccinated or SARS-CoV-2-infected mice or humans, *in vitro* microneutralization assays were performed similarly to what is described previously (13). Briefly, the mice or human sera were inactivated at 56°C for 30 min. Serum samples were serially diluted 3-fold starting from 1:10 dilution in Vero-E6-infection medium (DMEM+ 2% FBS+ 1% non-essential amino acids). The samples were incubated with optimized tissue culture infective dose 50 (TCID_50_), as described in the figure legends, of either WT- or MA-SARS-CoV-2 for 1 hour in an incubator at 37°C, 5% CO_2_ followed by incubation with pre-seeded Vero-E6 at 37°C for 48 hours. The plates were fixed in 4% formaldehyde at 4°C overnight. For TCID_50_ calculation, the virus stock was serially diluted 10-fold starting with 1:10 dilution and incubated on Vero-E6 cells for 48 hours followed by fixation in 4% Formaldehyde. The cells were washed with 1xPBS and permeabilized with 0.1% Triton X-100 in 1XPBS. The cells were washed again and blocked in 5% non-fat milk in 1xPBS+ 0.1% Tween-20 for 1 hour at room temperature. After blocking, the cells were incubated with anti-SARS-CoV-2 NP and anti-spike monoclonal antibodies, mixed in 1:1 ratio, for 1.5 hours at room temperature. The cells were washed in 1xPBS and incubated with 1:5000 diluted HRP-conjugated anti-mouse IgG secondary antibody for 1 hour at RT followed by a brief PBS wash. Finally, 100μl tetramethyl benzidine (TMB) substrate was added and incubated at RT until blue color appeared, and the reaction was terminated with 50μl 1M H_2_SO_4_. Absorbance was recorded at 450nm and 650nm and percentage reduction in infection was calculated as compared to negative control.

### Serum samples from human subjects

#### Ethics statement

The study protocols for the collection of clinical specimens from individuals with and without SARS-CoV-2 infection by the Personalized Virology Initiative were reviewed and approved by the Mount Sinai Hospital Institutional Review Board (IRB-16-00791; IRB-20-03374). All participants provided informed consent prior to collection of specimen and clinical information. All specimens were coded prior to processing.

#### Sample collection

A total of 34 sera were selected from study participants based on their SARS-CoV-2 spike enzyme linked immunosorbent assay (ELISA) antibody titer (negative [N=4] versus weak [N=8], moderate [N=11] or strong positive [N=11]). In addition, we included sera from six individuals that had received two doses of the Pfizer SARS-CoV-2 vaccine (V1-V6). Demographics and available metadata for each participant is summarized in Supplementary Table 1. Sera were heat-inactivated (56°C, 1 hour) and all experiments were conducted in a blinded manner.

## Results

### Serial passaging of SARS-CoV-2 in mice results in mouse-adapted SARS-CoV-2

The USA-WA1/2020-SARS-CoV-2 (termed WT-SARS-CoV-2) virus isolate was passaged eleven times in the lungs of various strains of mice as outlined in Fig. 1A. The virus was first allowed to adapt to murine ACE-2 receptor in immune-compromised mice with weak innate immune responses. To this end, the virus was consecutively passaged four times in IFNα/*λ* receptor knock-out mice in C57Bl6 genetic background, using 50 µl of lung homogenate from each infected mouse collected at three days post infection (DPI). The virus was then further passaged three times in BALB/c mice and four times in 129 mice. The 129 mice were chosen for mouse adaptation as they have been shown to be more susceptible to SARS-CoV, a virus that uses the same hACE-2 receptor as SARS-CoV-2. After eleven passages, the virus was plaque-purified and clonal virus stocks of the MA-SARS-CoV-2 were prepared in mACE-2 expressing Vero-E6 for further infection experiments. For comparative purposes, clonal virus stocks of the WT-SARS-CoV-2 were also generated using Vero-E6 cells. The consensus genomic sequence of the MA-SARS-CoV-2 was generated by sanger and deep sequencing methods and the sequence changes are summarized in Table 1. The MA-SARS-CoV-2 contained two amino acid mutations and one four amino acid insertion in the S, and one amino acid mutation in the M, N and ORF8 gene products of the virus (Fig. 1B). One of the amino-acid changes in the RBD of spike protein in the virus, N501Y, has previously been reported to be associated with mouse-adaptation of SARS-CoV-2 (21) and is predicted to increase binding to mACE-2 (14). Interestingly, the same N501Y mutation has recently been reported in newly emerging SARS-CoV-2 variants (20B/501Y.V1 strain) with potentially enhanced human transmission potential. Both WT- and MA-SARS-CoV-2 show similar growth kinetics in Vero E6 (Supplementary Fig. 2).

**Table 1:** Genomic changes in MA-SARS-CoV-2 (Passage P11) when compared with WT-SARS-CoV-2 (USA/WA1/2020-SARS-CoV-2).

| Segment | Nucleotide change | Amino acid change |
| --- | --- | --- |
| (S) Spike | A23063T<br>C23525T<br>22206 +12 insertion TAAGCTGAGAAG | N501Y<br>H655Y<br>+KLRS |
| (N) Nucleoprotein | T28853A | S194T |
| (M) Membrane | C26542T | T7I |
| ORF8 | T28144C | L84S |

**Fig. 1.**
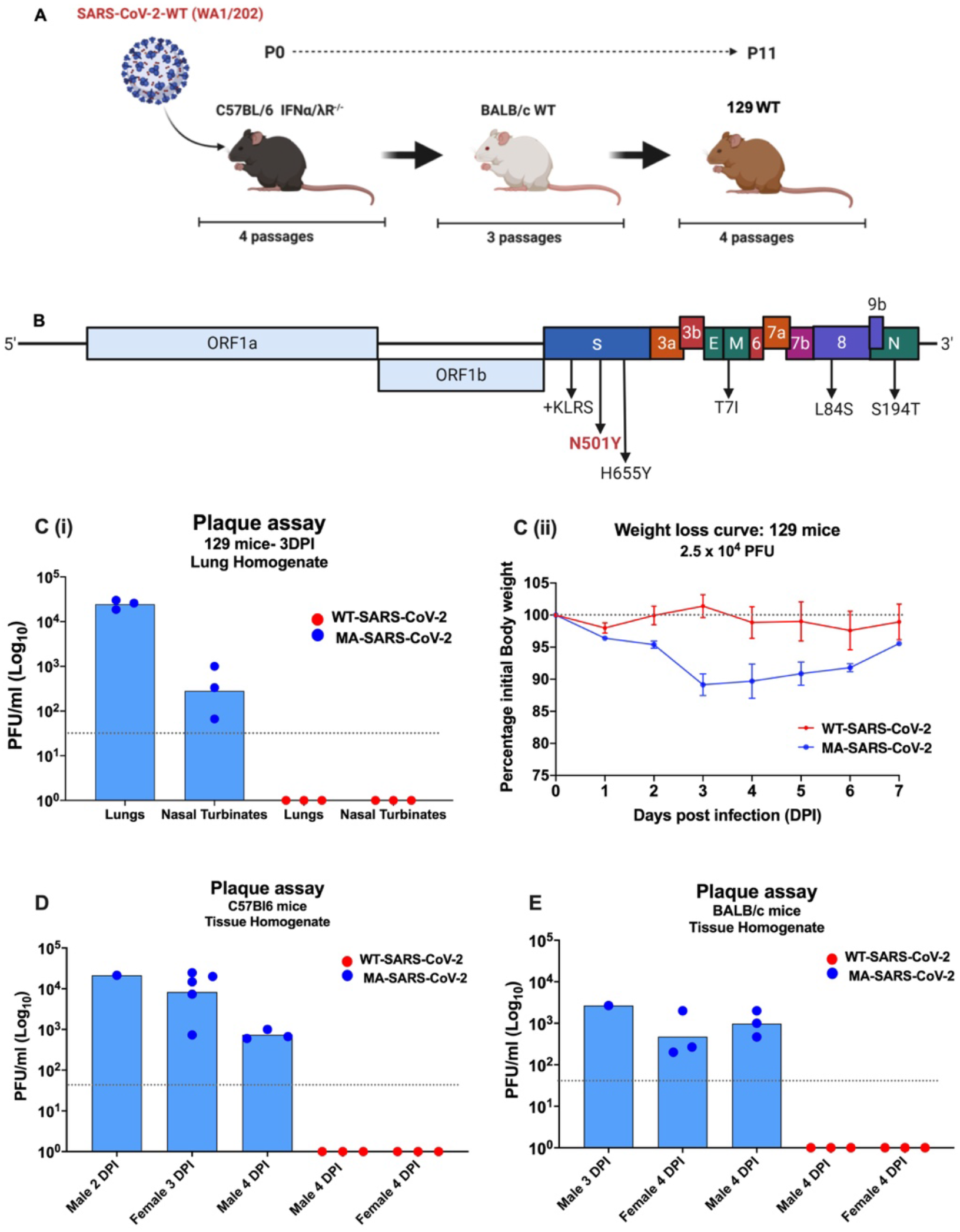
Characterization and mouse adaptation of MA-SARS-CoV-2. **(A)** The SARS-CoV-2 WA1/2020 Seattle strain was obtained from BEI Resources and serially passaged 11 times in mice of different genetic backgrounds. **(B)** Sequence and location of mutations identified in MA-SARS-CoV-2 when compared to WT-SARS-CoV-2. **(A)** Infection with 2.5 x 10^4^ PFU of the MA-SARS-CoV-2 resulted in detectable virus titers in lungs and nasal turbinate 3 DPI **(C-i)** as well as transient body weight loss in female 129 mice whereas the WT-SARS-CoV-2 did not **(C-ii). (D, E)** Infection with 2.5 x 10^4^ PFU of MA-SARS-CoV-2 but not WT-SARS-CoV-2, resulted in detectable lung virus titers harvested at different time points post-infection in all C57Bl6 **(D)** as well as BALB/c **(E)** mice irrespective of gender. Body weight loss was not observed in C57Bl6 or BALB/c mice strains upon infection with either of WT- or MA-SARS-CoV-2 (data not shown). Symbols represent geometric means; error bars represent standard deviation.

### MA-SARS-CoV-2 efficiently replicates in the lungs of different wild-type strains of mice and results in transient morbidity based on their genetic background

The pathogenicity of MA-SARS-CoV-2 was examined in terms of its infection and replication potential in laboratory mouse strains of different genetic backgrounds. While the WT-SARS-CoV-2 was unable to infect any of the laboratory strains of mice, MA-SARS-CoV-2 efficiently infected 129 (Fig. 1Ci), C57Bl6 (Fig. 1D) as well as BALB/c mice (Fig. 1E) with detectable virus titers in lungs and nasal turbinates of 129 mice and lungs of male and female C57Bl6 and BALB/c mice, obtained at different days post infection (DPI) as depicted in Fig. 1. MA-SARS-CoV-2 infection resulted in about 12% body weight loss in 129 mice (Fig. 1C-ii) while no weight loss was observed in infected C57Bl6 and BALB/c mice.

### MA-SARS-CoV-2 results in enhanced morbidity in mouse models of obesity, obesity-associated diabetes and advanced age

The successful establishment of a MA-SARS-CoV-2 allowed us to study risk factors previously associated with severe COVID-19 in humans. We focused on mouse models for obesity, diabetes and advanced age. Briefly, 6–8-week-old female C57Bl6 mice were fed with either control/low-fat (CD) or high-fat diet (HFD) for 14 weeks and the body weight changes were recorded over time. The mice on HFD gained almost twice the original weight and became obese (hence called obese mice) over time as compared to the mice on CD (also referred to as lean mice) (Fig. 2A). The diabetic status of the mice was determined by intraperitoneal glucose tolerance test (22), which suggested that the mice on HFD were diabetic while the mice on CD were not (Fig. 2B). The CD and HFD mice were then challenged with a low dose of MA-SARS-CoV-2 (1.7 ⨯ 10^3^ PFU/mice) and various organs including duodenum, heart, brain, kidney, lungs and pancreas were harvested five days-post-infection (Fig. 2C). Upon infection, the obese mice showed more morbidity, as reflected in higher body weight loss over five days-post-infection as well as on higher lung viral titers when compared to the lean mice (Fig. 2D-i and D-ii). We also tested our MA-SARS-CoV-2 on 6-8-week-old and 52-week-old female C57Bl6 mice, also referred to as young and old mice, respectively. With the same low viral challenge dose (1.7 ⨯ 10^3^ PFU/mice), the 52-week-old mice showed more morbidity as reflected by body weight loss over five days-post-infection when compared to young mice. The 52-week-old mice also showed physical symptoms of distress including hunched back and difficulty and reluctance in movement while the 6-8-week-old mice resumed normal activity. Additionally, the viral lung titers were also found to be higher in 52-week-old mice as compared to 6-8-week-old mice. five days-post-infection. Our data summarizes that both obesity/diabetes and advanced age in mice result in higher morbidity during SARS-CoV-2 infection. Although we did not find detectable virus titers in other organs with low virus challenge in obese or aged mice, this does not exclude possible extrapulmonary viral replication at other time points or with a higher dose of virus challenge.

**Fig. 2.**
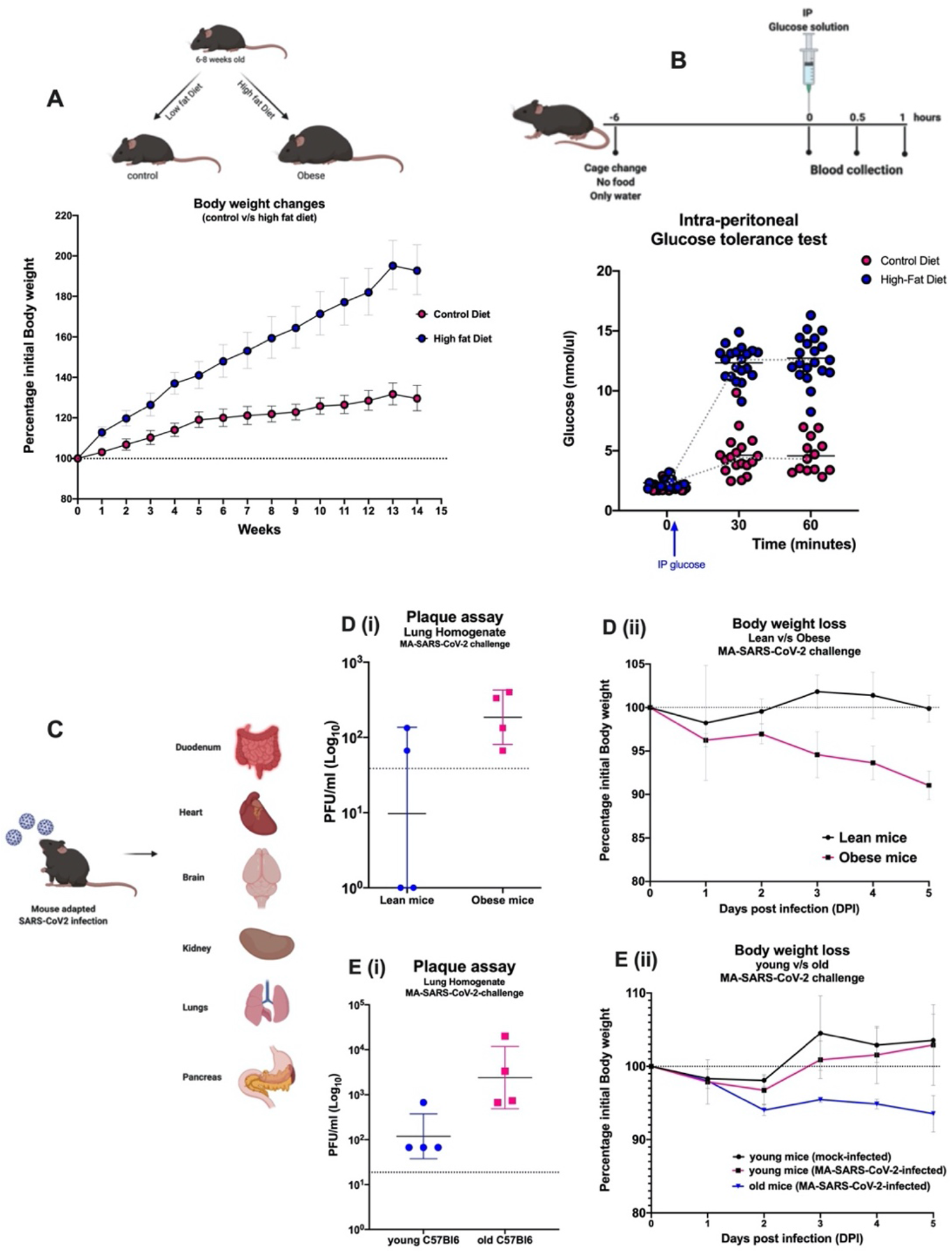
Age and obesity as risk factors for severity associated with MA-SARS-CoV-2. **(A)** 6-8-week-old C57Bl6 mice were fed with sucrose-matched high fat or control diet for up to 14 weeks and the body weight changes were recorded. **(B)** Mice on control or high-fat diet were fasted for six hours followed by an intraperitoneal injection of dextrose solution at 2g/kg body weight. Blood was drawn at different time points by submandibular bleed and blood glucose levels were determined by Glucose assay. (C) Diagrammatic representation of intranasal infection with 1700 PFU/mouse of SARS-CoV-2 variant (MA-SARS-CoV-2) followed by harvest of various organs to assess virus replication. **(D, E)** the body weight changes were monitored post-infection until the harvest. Higher lung virus titers were observed in obese mice **(D-i)** accompanied with noticeable loss in body weight **(D-ii)** as compared to lean/control diet mice. Similarly, the lung virus titers **(E-i)** and body weight loss **(E-ii)** were found to be higher in 52-week-old mice as compared to 6-8-weeks young/control mice, five days-post-infection. No virus titers were found in other organs harvested five days post infection in both experiments. Symbols represent geometric means; error bars represent standard deviation.

### N501Y mutation does not affect SARS-CoV-2 neutralization by mouse convalescent and post vaccination serum

The WT-SARS-CoV-2 and MA-SARS-CoV-2-infected 129 mice were further tested for the presence of SARS-CoV-2-specific neutralizing antibodies against both viral strains. As shown in Fig. 3, the MA-SARS-CoV-2 post-challenge sera were able to neutralize both WT-SARS-CoV-2 (Fig. 3A-i and A-ii) as well as MA-SARS-CoV-2 (Fig. 3B-i and B-ii) in *in vitro* microneutralization assays. Additionally, no neutralizing antibody titers were observed in the sera of 129 mice infected with WT-SARS-CoV-2 against both strains of viruses, which is in line with WT-SARS-CoV-2 not being able to infect or replicate efficiently in 129 mice lungs and therefore, negligible immune responses were induced in the mice. Besides post-challenge studies, we also performed *in vitro* microneutralization assays using 3-week post-vaccination sera from BALB/c mice that received an adjuvanted recombinant SARS-CoV-2 S protein vaccine (from a vaccination study reported in our recent prepublication (15) that is currently under revision). The adjuvanted-spike vaccination sera were found to be neutralizing to the same extent against both strains of the virus, irrespective of the N501Y mutation (Fig. 3C and 3D).

**Fig. 3.**
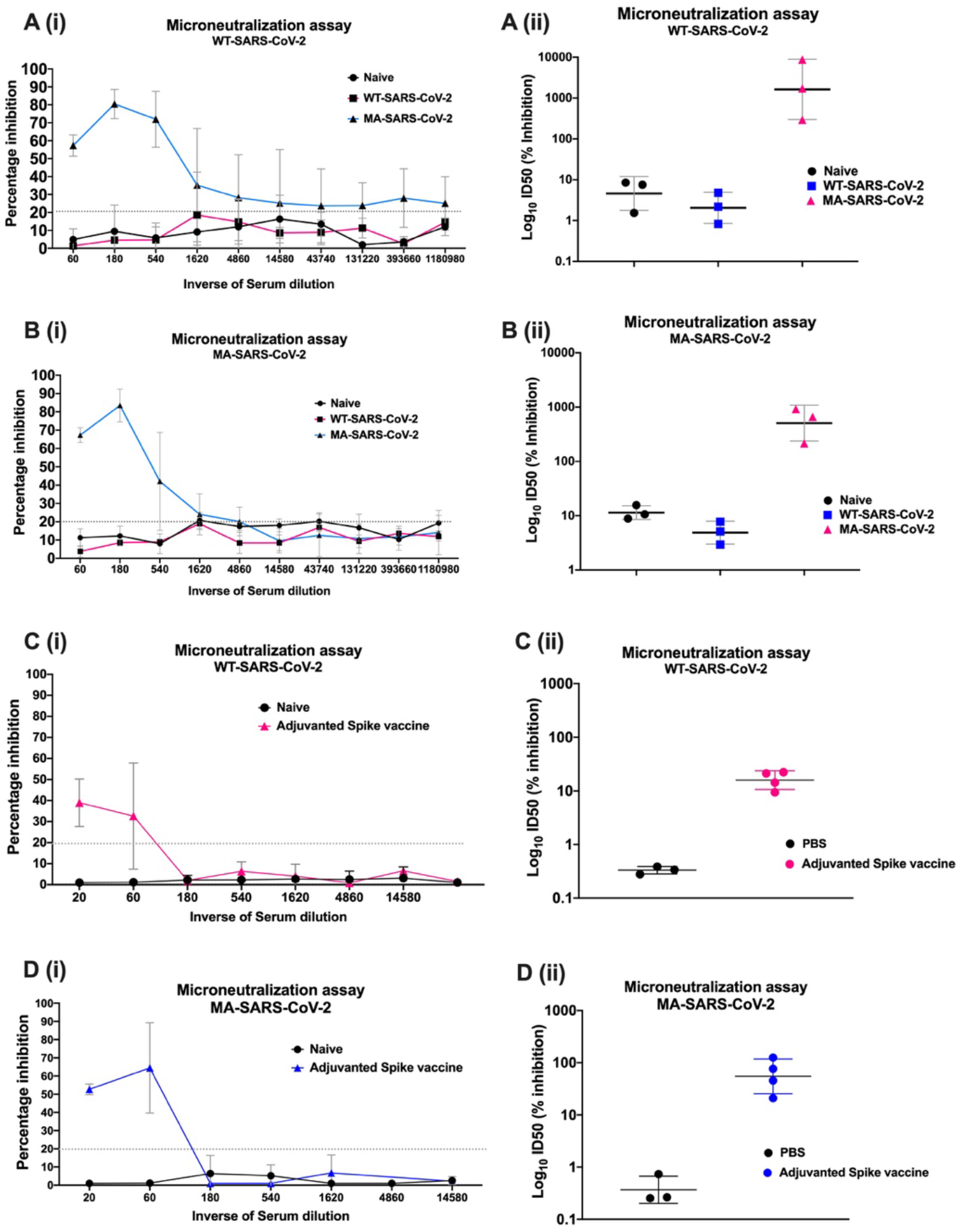
Sera from mice vaccinated with WT SARS-CoV-2-S protein or pre-exposed to MA-SARS-CoV-2 can effectively neutralize both WT- and MA-SARS-CoV-2 virus. **(A, B)** 129 mice were mock-infected or infected with 2.5 x 10^4^ PFU/mouse of either of WT- or MA-SARS-CoV-2. Blood was drawn from mice 3-weeks post-infection and microneutralization assays were performed against WT-SARS-CoV-2 **(A-i and A-ii)** or MA-SARS-CoV-2 **(B-i and B-ii)** using 200TCID_50_ of each virus. Sera from mice pre-exposed to MA-SARS-CoV-2 were able to neutralize both strains of virus. **(C, D)** Sera from BALB/c mice, non-vaccinated or vaccinated with adjuvanted-SARS-CoV-2-recombinant S-protein, were tested for presence of neutralizing antibodies by *in vitro* microneutralization assays with WT-SARS-CoV-2 **(C-i and C-ii)** or MA-SARS-CoV-2 **(D-i and D-ii)**. Sera from mice vaccinated with adjuvanted-spike protein was able to neutralize both strains of virus. Sera from non-vaccinated mice were used as control in the experiment. Symbols represent geometric means; error bars represent standard deviation.

### N501Y mutation does not affect SARS-CoV-2 neutralization by human convalescent and post vaccination serum

The comparison of post-vaccination and post-challenge sera from mice showed that both WT- and MA-SARS-CoV-2 strains were neutralized to the same extent indicating that the N501Y substitution in the receptor binding domain of Spike does not mediate antibody escape. To further validate these observations, we next performed similar microneutralization assays using sera from study participants with or without SARS-CoV-2 immune responses. We included samples from individuals who had recovered from natural infection or had received the Pfizer vaccine (see supplementary table 1). Sera were tested for neutralizing potential against both WT- as well as MA-SARS-CoV-2 strains. Post-vaccination sera had neutralizing antibody titers that were similar to the highest neutralization titers observed in convalescent sera (Fig. 4). Moreover, microneutralization titers against both WT- and MA-SARS-CoV-2 were comparable. Overall, our study shows that the N501Y mutation in the RBD domain of the SARS-CoV-2 spike protein does not compromise the neutralization potential of this virus by convalescent and post-vaccination human sera.

**Fig. 4.**
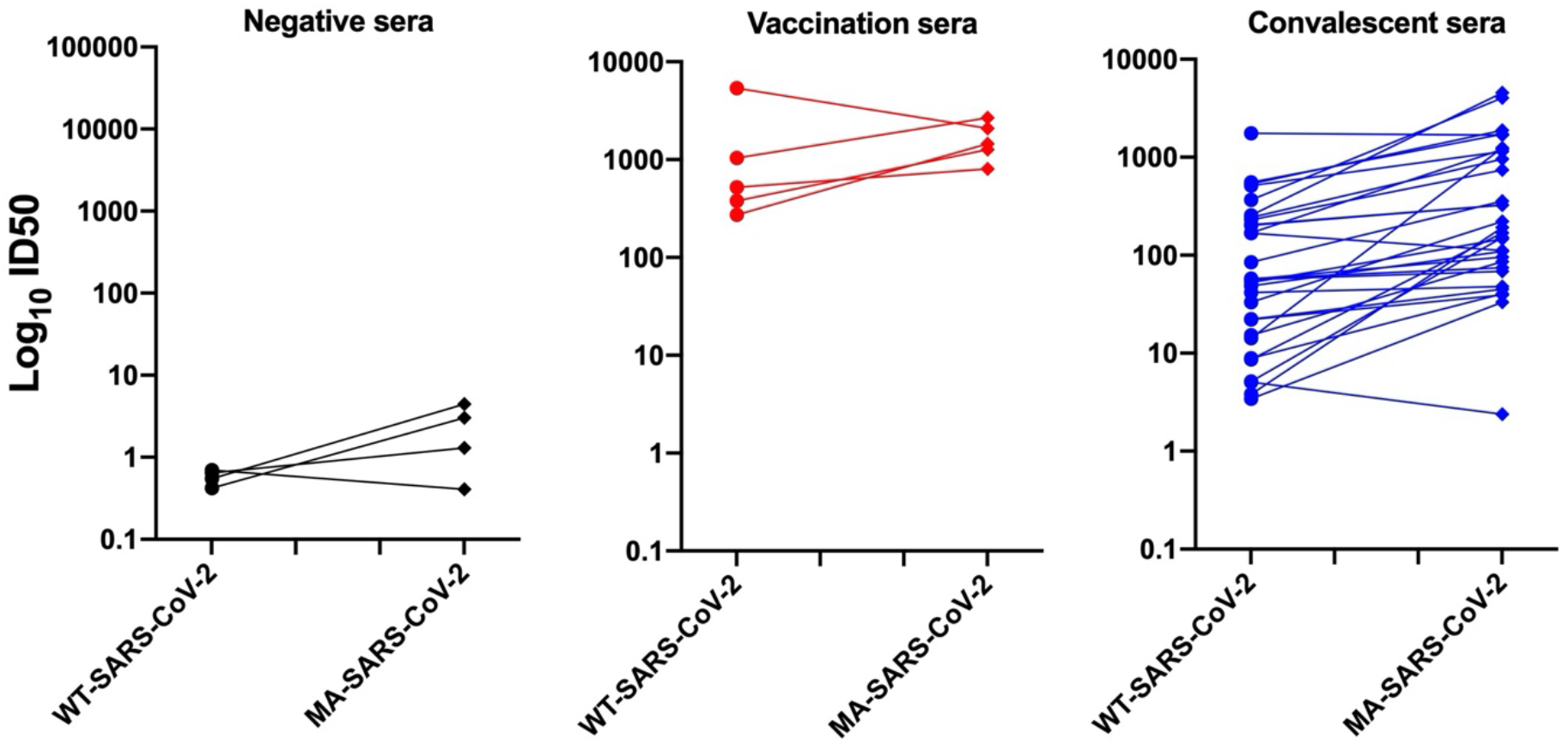
Post-vaccination or convalescent human sera neutralize both WT- and N501Y MA-SARS-CoV-2 viruses. Post-vaccination or post-infection human sera were analyzed for virus neutralization by *in vitro* microneutralization assays using 450TCID_50_ of either WT-SARS-CoV-2 or MA-SARS-CoV-2 and the ID50 (inhibition) values were calculated and compared. Each symbol represents ID50 (inhibition) value calculated for a human serum sample. Both post-vaccination sera as well as convalescent sera neutralize both strains of virus. Sera from sero-negative individuals were used as negative control for the experiment.

## Discussion

There is an urgent need for animal models to test efficacies of antiviral interventions and to study underlying risk factors associated with enhanced illness during COVID-19. We -adapted the USA-WA1/2020 strain to mice by serial passaging the virus in mouse lungs. This resulted in acquisition of mutations that allow the virus to efficiently replicate to detectable titers in the lungs of mice. Mouse adaptation did not impact growth kinetics of the virus on Vero E6 cells, which suggests that mutations associated with mouse-adaptation broadened receptor specificity rather than changed receptor specificity from human (or primate) ACE-2 to murine ACE-2. We observed multiple mutations associated in different viral genes. The MA-SARS-CoV-2 has acquired an asparagine to tyrosine mutation at position 501 in the spike RBD (N501Y). The same mutation has recently been described by Gu *et al*. (2020) for mouse-adaptation of SARS-CoV-2 following a similar strategy with aged BALB/c mice (16). *In silico* analysis also predicted that the N501Y mutation would result in enhanced RBD affinity for the mACE-2 receptor (14). We therefore concluded that this mutation is important for mouse adaptation. We also observed a four amino acid insertion in the spike protein (KLRS). This insertion has been described in SARS-CoV-2 viruses recovered from deer mice upon experimental infection with the same USA-WA1/2020 strain (17). Minority variants with this insertion were already present in the original SARS-CoV-2 virus obtained from BEI Resources at low frequency and seems also to be enriched after passaging in Vero E6 cells (data not shown) and mice (this study). Interestingly, cats have recently been identified as an animal species sensitive to SARS-CoV-2 infection (18) and SARS-CoV-2 with the KLRS insertion was also enriched in lung and GI tract tissues as well as nasal, oropharyngeal and rectal swabs from infected cats upon experimental infection (supplementary Fig. 3). Therefore, it is likely that the KLRS insertion increases viral replication in multiple hosts, not only in mice. The effect of presence of the KLRS insertion is currently being investigated. We also observed mutations outside of the spike protein of the MA-SARS-CoV-2. As has been shown for influenza virus and SARS-CoV before, multiple viral gene products may contribute to virulence, either individually or by interaction with each other (19–21). At this point it is still unclear if the mutations outside of spike protein contribute to increased replication and virulence in mice or if they are passenger mutations. Introduction of the observed mutations in a recombinant virus approach for SARS-CoV-2 infectious clone is needed to investigate the individual or synergistic contributions to replication and virulence of these mutations.

Interestingly, the N501Y mutation in the RBD of the S protein is also found in the newly emerging 20B/501Y.V1 strain that was first detected in the UK but now is circulating worldwide with high transmissibility. The N501Y mutation is the only mutation in the RBD of the S protein of the UK-origin 20B/501Y.V1 strains, and therefore, might impact its antigenicity. The vaccines that are currently being rolled out have an N at position 501. Viral escape from neutralizing antibodies has been shown *in vitro* using pseudoviruses (22) and there are strong concerns that monoclonal antibody therapies and vaccines that become currently available may have reduced efficacy against circulating strains with the N501Y mutation. We showed that convalescent and post-vaccination sera obtained from both mice and humans can still potently neutralize the MA-SARS-CoV-2 variant with N501Y mutation in its RBD. This strongly suggests that humoral immune responses induced by the SARS-CoV-2 vaccines that are currently entering the market will be protective against the newly emerging 20B/501Y.V1 strain. This is especially important for people with comorbidities that are at risk for severe illness during COVID-19 and therefore, are prioritized for vaccination. Comorbidities include advanced age, type 2 diabetes mellitus and obesity. Obesity was a predictor for mortality among intensive care unit-admitted patients with severe COVID-19, and put patients at higher risk for hypoxemia (6). Diabetes increases mortality among COVID-19 patients (3). In this work we present two mouse models for studying enhanced illness during the outcome of SARS-CoV-2 infection in the context of these comorbidities with the MA-SARS-CoV-2 that has the N501Y mutation. This mouse model does not rely on transgenic expression or adenoviral transduction of hACE-2. Therefore, it is suitable to study the effect of ACE inhibitors and angiotensin II type I-receptor blockers that are used to treat diabetics and can result in upregulation of hACE-2, thereby potentially sensitizing patients for severe SARS-CoV-2 infection (23,24).

## Data Availability

Data is available upon request.

## Acknowledgements

PVI study group: Dr. G. Kleiner, Dr. M. Saksena, K. Srivastava, C. Gleason, C. M. Bermúdez-González, K. Beach, K. Russo, L. Sominsky, E. Ferreri, R. Chernet, L. Eaker, A. Salimbangon, D. Jurczyszak, H. Alshammary, W. Mendez, A. Amoako, S. Fabre, M. Awawda, A. Shin

We also thank Dr. Randy Albrecht for support with the BSL3 facility and procedures at the ISMMS as well as Richard Cadagan for excellent technical assistance.

## Funding

This work was partially supported by the NIAID Centers of Excellence for Influenza Research and Surveillance (CEIRS) contract HHSN272201400008C (to A.G.-S., F.K. and V.S.) and contract HHSN 272201400006C (to J.A.R.), Collaborative Influenza Vaccine Innovation Centers (CIVIC) contract 75N93019C00051 (to A.G.-S., F.K. and V.S.), and the generous support of the JPB foundation (to A.G.-S., F.K. and V.S.), the Open Philanthropy Project (#2020-215611) (to A.G.-S., F.K., V.S.) and other philanthropic donations. This research was partly funded by NIH AI150355 grant to LCFM, by CRIP (Center for Research for Influenza Pathogenesis), a NIAID supported Center of Excellence for Influenza Research and Surveillance (CEIRS, contract # HHSN272201400008C), by the Defense Advanced Research Projects Agency (HR0011-19-2-0020), by supplements to NIAID grant U19AI135972 and DoD grant W81XWH-20-1-0270, by NCI grant U54CA260560, by a Mercatus Center Fast Grant, by the generous support of JPB Foundation, the Open Philanthropy Project (research grant 2020-215611 (5384); by anonymous donors to A.G.-S.. Funding for this study was provided through grants from the National Bio and Agro-Defense Facility (NBAF) Transition Funds from the State of Kansas, and DHS CEEZAD (HSHQDC16-A-B0006) to J.A.R. B.G.D.G acknowledges funding from the European Research Council (ERC) under the European Union’s Horizon 2020 research and innovation program (grant N 817938). S.Y. is supported by The Swiss National Science Foundation (Early Postdoc Mobility Grant Project Id: P2GEP3_184202).

## Author contributions

R.R., S.J., A.Cupic., C.M.R., L.C.F.M., T.K., S.Y., A.Choi., I.M., S.A., D.S., D.A.M., C.D.M., V.B., M.S. designed and/or performed experiments. The PVI study group recruited, consented, enrolled participants, collected clinical data and obtained, processed and banked clinical specimen used in this study. R.R., S.J., C.M.R., S.Y., A. Choi, I.M., S.A., D.A.M., C.D.M., V.B., V.S., B.G.D.G. and M.S. analysed data. L.C.F.M., L.M., J.D.V., F.K., B.G.D.G. generated critical reagents. J.A.R., F.K., B.G.D.G, V.S., A.G.-S. and M.S. oversaw the conception and design of the experiments. R.R., S.J., F.K., V.S., A.G.-S. and M.S. wrote the manuscript or substantively revised it.

## Conflicts of interest

The A.G.-S. laboratory has received research support from Pfizer, Senhwa Biosciences, Kenall Manufacturing, Avimex, Johnson & Johnson, Dynavax, 7Hills Pharma, ImmunityBio and Nanocomposix. Dr. Adolfo García-Sastre has consulting agreements for the following companies involving cash and/or stock: Vivaldi Biosciences, Contrafect, 7Hills Pharma, Avimex, Vaxalto, Pagoda, Accurius and Esperovax. Mount Sinai has licensed SARS-CoV-2 serological assays to commercial entities and has filed for patent protection for serological assays as well as SARS-CoV-2 vaccines. F.K. is listed as inventors on the pending patent applications. The F.K. laboratory has received research support from GSK, Dynavax and Pfizer. F.K. has in the past received consulting fees from Curevac, Merck, Pfizer and Seqirus. A provisional patent application on a “A novel 4 Amino Acid Insertion into the Spike Protein of SARS-CoV-2” was submitted by KSU in July 2020 with C.D.M., D.A.M and J.A.R listed as inventors.

---

***Supplementary 1***: *Stably expressing mACE-2 Vero E6 cell line*

**Supplementary Fig. 1.**
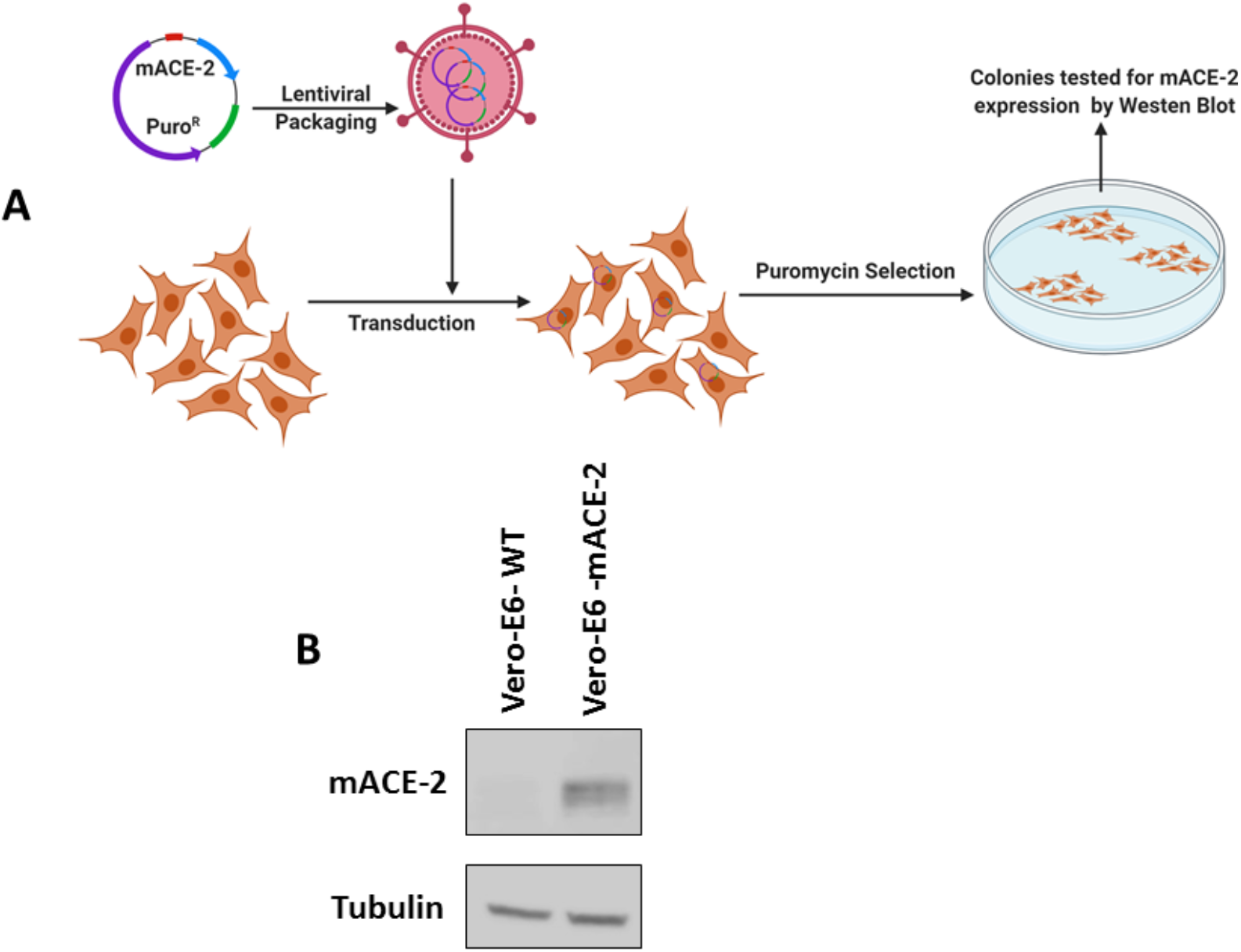
Establishing mACE-2 Vero-E6 cells. **(A)** Strategy: Vero-E6 cells were transduced with a lentiviral vector expressing mACE-2 and a puromycin resistance gene. Cells were selected for mACE-2 expression by puromycin selection. **(B)** Expression in the selected polyclonal population was confirmed by Western blot.

***Supplementary 2***: *Growth kinetics of WT and MA-SARS-CoV-2 on Vero-E6 cells*

**Supplementary Fig. 2.**
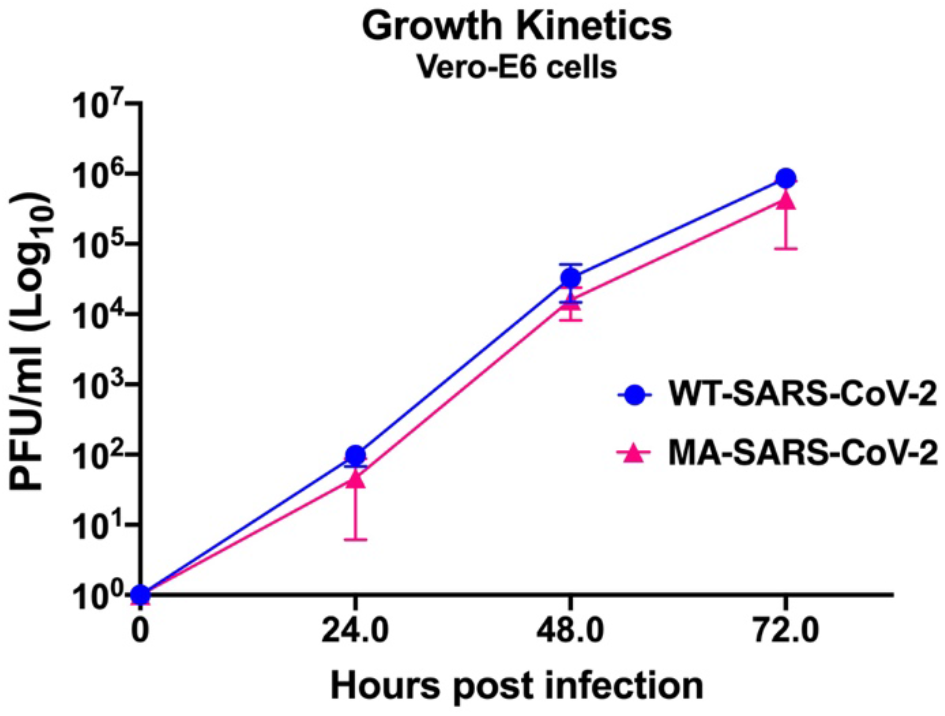
Comparison of growth kinetics of WT-SARS-CoV2 and MA-SARS-CoV-2. Vero-E6 cells were infected with equal PFUs of WT-SARS-CoV-2 or MA-SARS-CoV-2 and supernatant media was collected at different time points. The virus replication was titrated by plaque assay. No major difference was observed in growth kinetics of WT and MA-SARS-CoV-2 *in-vitro*. Symbols represent means, error bars represent standard error.

***Supplementary 3***: *KLRS insertion in SARS-CoV-2 after infection in cats*

**Supplementary Fig. 3:**
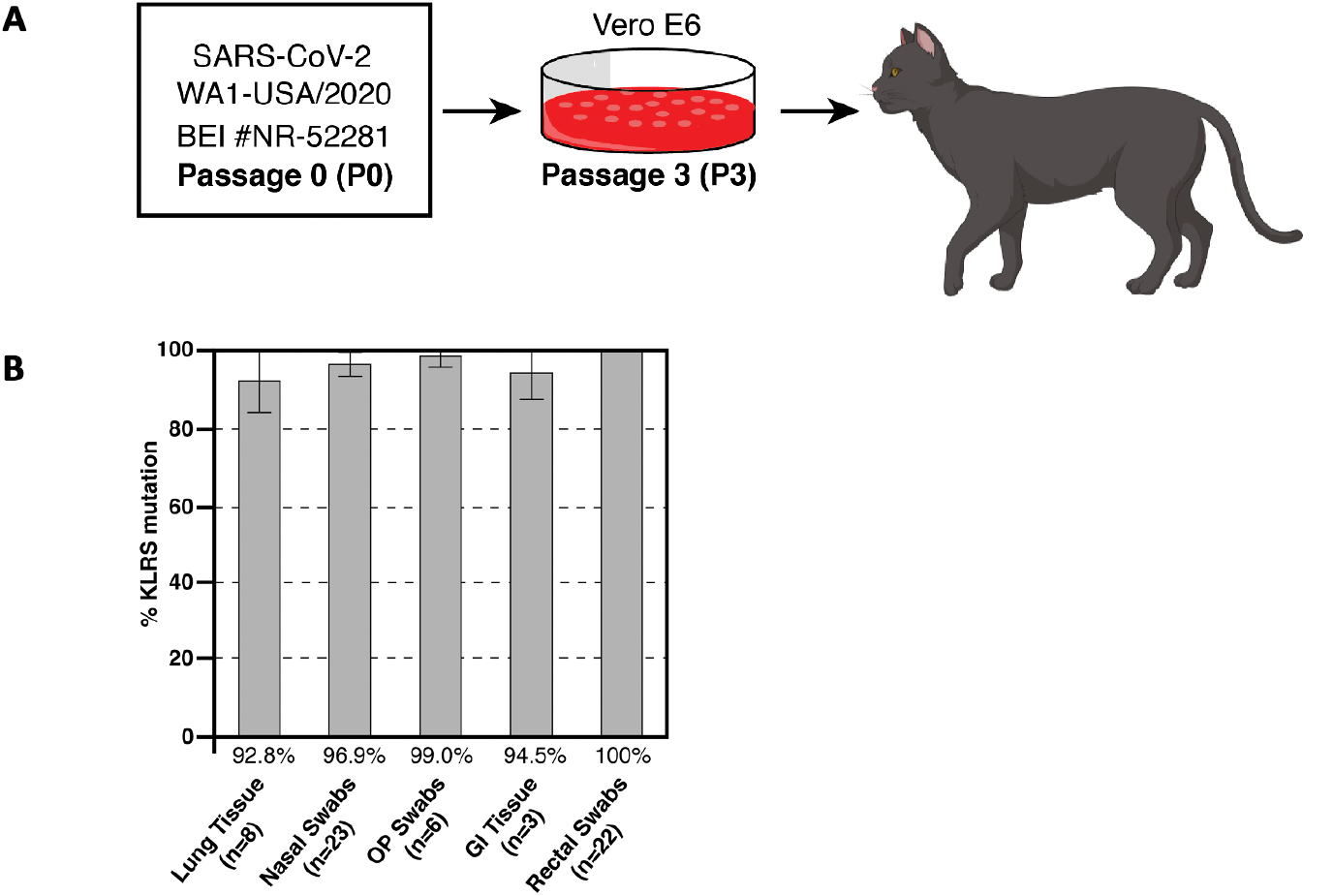
Increased prevalence of KLRS insertion in SARS-CoV-2-infected cats. (A) The SARS-CoV-2 WA1-USA/2020 strain from BEI was passaged three times in Vero E6 cells before being used to infect six cats intranasally and orally with a 10^6^ TCID_50_ dose of virus that were subsequently exposed to 2 sentinel contact cats that also became infected. Nasal, oropharyngeal (OP), and rectal swabs were collected from cats on 2 to 7 days post-challenge (DPC) and lung/Gastrointestinal (GI) tissues were collected on 4-7 DPC. RNA was extracted, sequenced, and analyzed to determine the relative percentage of the KLRS mutation in various clinical samples from cats. (B) Chart showing that the prevalence of the KLRS mutation increases in cats with 90% −100% prevalence in various swabs or tissues.

**Supplementary table 1:** Description of serum samples obtained from human subjects

| SERUM |  |  |  |  |
| --- | --- | --- | --- | --- |
| Seropositive, vaccine | Spike IgG response | Sex | Age group (yrs) | Days post 1 vaccine dose (Pfizer) |
| V1 | Strong positive | F | >60 | 68 |
| V2 | Strong positive | M | 30-40 | 47 |
| V3 | Strong positive | F | 50-60 | 47 |
| V4 | Strong positive | M | >60 | 48 |
| V5 | Strong positive | F | 40-50 | 49 |
| V6 | Strong positive | F | 30-40 | 48 |
| Seropositive, infection | Spike IgG response | Sex | Age group (yrs) | Days post onset of symptoms |
| P1 | Weak positive | M | 20-29 | 260 |
| P2 | Weak positive | M | 50-59 | NA |
| P3 | Weak positive | F | 30-39 | 111 |
| P4 | Weak positive | F | 30-39 | 221 |
| P5 | Weak positive | F | 30-39 | 254 |
| P6 | Weak positive | F | 20-29 | 247 |
| P7 | Weak positive | M | 30-39 | 220 |
| P8 | Weak positive | F | 20-29 | Asymptomatic |
| P9 | Moderate positive | M | 30-39 | NA |
| P10 | Moderate positive | F | 30-39 | 197 |
| P11 | Moderate positive | F | 50-59 | Asymptomatic |
| P12 | Moderate positive | F | 30-39 | Asymptomatic |
| P13 | Moderate positive | M | 30-39 | 234 |
| P14 | Moderate positive | F | 20-29 | 273 |
| P15 | Moderate positive | M | 30-39 | Asymptomatic |
| P16 | Moderate positive | F | 20-29 | 258 |
| P17 | Moderate positive | F | 20-29 | 246 |
| P18 | Moderate positive | M | 20-29 | Asymptomatic |
| P19 | Moderate positive | F | 50-59 | 204 |
| P20 | Strong positive | F | 50-59 | NA |
| P21 | Strong positive | F | 30-39 | 245 |
| P22 | Strong positive | M | NA | 170 |
| P23 | Strong positive | F | >60 | Asymptomatic |
| P24 | Strong positive | F | 40-49 | NA |
| P25 | Strong positive | F | 50-59 | 191 |
| P26 | Strong positive | F | 30-39 | NA |
| P27 | Strong positive | F | 50-59 | 113 |
| P28 | Strong positive | M | >60 | Asymptomatic |
| P29 | Strong positive | M | 18-19 | 218 |
| P30 | Strong positive | M | 50-59 | 219 |

|  | Seronegative,<br>post pandemic | Spike IgG response | Sex | Age group<br>(yrs) | Days from last negative<br>serology |
| --- | --- | --- | --- | --- | --- |
|  | N1 | Negative | F | 40-50 | 23 |
|  | N2 | Negative | F | 20-29 | 24 |
|  | N3 | Negative | F | 20-29 | 23 |
|  | N4 | Negative | F | 30-35 | 22 |

## Notes

### Clinical Trial

NA

